# Muévete conCiencia: Study protocol for a randomized controlled trial of dual-task exercise, Tai Chi, and cognitive training on executive functions and stress in university students

**DOI:** 10.64898/2026.03.20.26348951

**Authors:** Montserrat Rodríguez-Vera, Carolina Pinto, Claudio Báez, Claudia Llanos, Axel Koch, Daniel Reyes-Molina, Daniel Peña-Oyarzún, Samira Rostami, Iria de la Osa Subtil, Cristina Perdomo-Delgado

## Abstract

The transition to higher education is characterized by increased academic demands and psychosocial stress, which may negatively affect cognitive functioning and student well-being. Executive functions (working memory, inhibitory control, and cognitive flexibility) are critical for academic adaptation and can be enhanced through structured interventions. Physical exercise, mind–body practices, and cognitive training have demonstrated potential benefits for executive functioning and stress reduction; however, few randomized controlled trials have directly compared interventions with different physical and cognitive demands in university students, particularly in Latin America. In addition, most studies have relied on self-report measures and physiological stress biomarkers such as salivary cortisol.

This protocol describes a three-arm, parallel-group randomized controlled trial designed to evaluate the effects of a 12-week intervention on executive functions and stress in first-year university students. The study will recruit 160 first-year health-science students aged 18–25 years. Participants will be randomly assigned (1:1:1), using block randomization stratified by sex, to one of three interventions delivered twice weekly (24 sessions of 60 minutes): (1) moderate-to-vigorous motor–cognitive dual-task exercise (DT); (2) low-to-moderate intensity Tai Chi (TC); or (3) supervised digital cognitive training (CT) using structured graphomotor tasks.

Primary outcomes include executive functions assessed through standardized neuropsychological measures. Secondary outcomes include stress will be evaluated using the Academic Stress Inventory, Depression Anxiety Stress Scales and salivary cortisol collected in the morning using passive drool and analyzed by competitive ELISA.Other outcomes include physical activity levels, anthropometric and body composition measures, and handgrip strength. Data will be analyzed following an intention-to-treat approach using repeated-measures models, with multiple imputation for missing data.

The study has received institutional ethics and biosafety approval. Trial registration: ClinicalTrials.gov Identifier: NCT07443904 27/02/26.

## Introduction

Entering higher education is a stage marked by major personal, academic, and social challenges for university students. In Chile, 26.4% of first-year students drop out [1], which is a concern for students, their families, and higher education institutions [2]. This phenomenon has driven the development of programs aimed at promoting student adjustment and retention, an objective shared by researchers, institutions, governments, and international organizations [3–6].

Among the factors predicting university adjustment, cognitive resources have been shown to be fundamental [7,8]. In particular, executive functions (EF)—higher-order processes mediated by the frontal lobe—are significantly associated with academic performance (r = .27) [9] and support self-regulated learning and problem solving [10–12]. EF comprises three core components: working memory, inhibitory control, and cognitive flexibility [13]. Studies have shown that students with stronger executive functioning achieve better academic outcomes and progress more successfully [14,15]. EF can be trained and improved through structured practice, with positive effects reported even in short-duration, high-intensity interventions [16]. Because EF development continues into early adulthood, higher education represents a particularly relevant context for EF stimulation [14].

Stress is a risk factor for academic progress during the university period [17,18]. Sustained exposure to high stress levels has been associated with cognitive alterations in working memory and cognitive flexibility, as well as sleep and health disturbances [19,20]. This is especially relevant for first-year university students, who face multiple environmental changes, new routines, and new learning demands [21].

Exposure to psychosocial stressors triggers activation activation of the sympathetic–adrenal–medullary system and the hypothalamic–pituitary–adrenal axis, increasing cortisol release [22]. This adrenal hormone shows a strong circadian rhythm, with high morning levels and a decline toward night [23]. A daily cortisol slope with a typically steep decline is associated with better physical and psychosocial health [23]. This biomarker has been linked to a range of stress-related disorders [24]. Evidence indicates that cortisol assessment using saliva and blood are valid methods for capturing short-term cortisol increases [25].

In addition, university environments are characterized by high academic demands and long hours of study, creating contexts of low physical activity [26,27]. The World Health Organization reports that one third of adults do not meet recommendations of 150 minutes per week of moderate physical activity [28]. Physical inactivity is associated with increased risk of cardiovascular disease, cognitive decline, and mood disorders, whereas regular physical activity is linked to better physical and mental health and greater well-being [29–31].

Regular physical exercise has demonstrated cognitive benefits, including improvements in attention and EF, as well as reductions in stress, anxiety, and depressive symptoms in university students [10,30,32]. These effects are modulated by both exercise intensity and structure, with benefits observed for moderate-to-vigorous aerobic programs [33] and for programs that incorporate dual-task demands [34]. Dual-tasks (DT) are defined as the simultaneous or alternating performance of two or more motor and cognitive tasks [35]. This training involves both automatic processing (fast and autonomous) and controlled processing (slower, involving working memory and attentional control) [36]. In athletes, negative effects have been reported in acute responses, but better outcomes have been observed over the long term [36]. Accordingly, DT has been proposed as a preventive intervention to improve EF, with potential direct or indirect benefits for cognitive capacity, academic performance, and mental health in young people [35].

A contemporary modality is mind–body physical training, which has shown positive effects on physical, mental, and cognitive health [37]. These practices integrate body awareness and mindfulness during movement and include disciplines such as Tai Chi (TC), yoga, and qigong [38,39]. TC has been used as a low-to-moderate intensity exercise incorporating a sequence of postures linked by smooth movements, showing effects on physical function, psychological health, quality of life, sleep quality, self-efficacy, stress, anxiety, depression, and mood in university students [40]. Li et al. [41] reported that TC significantly improves prefrontal lobe activation, neuroplasticity, and reaction speed, evidenced in the alpha band of electroencephalography. The most commonly used style is Yang-style TC, which consists of simplified movements and postures to facilitate efficient learning and wide dissemination [40]. TC has been applied both in older adults and in university populations [40]. It integrates martial arts and meditation, with the advantages of being easy to learn, safe, and effective [41]. A systematic review by Kraft et al.[42] found that TC reduces stress, with a durable effect of small-to-moderate magnitude [42], and validated these effects in healthy populations across age groups (young people, adults, and older adults). Despite these findings, research remains limited by a lack of prospective, rigorous, high-quality studies to define optimal TC frequency and duration, constraining broader implementation [40,42].

Cognitive training (CT), and particularly digital cognitive training, constitutes a key component in strengthening EF, understood as a set of high-level mental abilities that include planning, working memory, inhibitory control, cognitive flexibility, and decision-making [43]. The use of digital platforms and applications allows for the presentation of adaptive tasks, immediate feedback, and precise control of cognitive load—features that are especially relevant in university populations, which are characterized by a high level of digital literacy. These tools facilitate personalized and scalable interventions, increasing adherence and the ecological validity of CT programs [44]. Evidence suggests that digital cognitive training, when integrated with physical exercise, enhances neuroplasticity and the functioning of frontoparietal networks involved in executive control [45,46]. Likewise, physical exercise improves the efficiency of attentional processes and working memory, creating a neurobiological environment that is favorable for consolidating the effects of digital CT [46].

Despite the growing evidence on the benefits of physical exercise and mind–body interventions for health and cognitive functioning, important knowledge gaps remain for university populations. Randomized controlled trials directly comparing exercise modalities with different physical and cognitive demand profiles are scarce, particularly in higher education students [10,47]. Moreover, few studies have incorporated physiological biomarkers such as cortisol to better understand underlying stress mechanisms [20]. In Latin America, and in Chile in particular, longitudinal and experimental research on physical exercise, EF, and stress in university students is still incipient [48]. These gaps justify the development of a randomized trial comparing physical exercise and mind–body interventions with different mechanisms of action, integrating assessments of EF, psychological stress, cortisol, and physical fitness in first-year students, to inform effective health promotion strategies and academic adaptation in the Chilean university context.

In summary, DT, TC, and CT represent promising strategies to promote cognitive strengthening and reduce stress among university students. DT, particularly in higher-intensity formats, offers an efficient intervention integrating simultaneous motor and cognitive demands that may enhance EF training. TC is a low-to-moderate intensity mind–body modality that may support emotional self-regulation, mindfulness, and mind–body integration. CS provides focused and structured CT with the potential to directly impact EF. Although emerging evidence supports the individual benefits of these approaches, clinical trials comparing their differential effects on EF, stress, and cortisol in university populations especially in Chile remain scarce.

### General objective

Evaluate between group differences in pre-post intervention in EF and stress across the three interventions arms DT, TC, CT in university students.

### Specific objectives

The first objective is to determine levels of EF, psychological stress (self-report and salivary cortisol), and physical fitness (physical exercise level and anthropometric variables) in university students before and after the 12-week intervention. The second objective is to compare pre–post changes in EF, psychological stress, and physical fitness across the DT, TC, and CT groups following the 12-week intervention.

### Hypotheses

#### Theoretical Hypothesis 1 – EF

It is expected that the DT intervention will produce greater improvements in EF compared with the TC group and the CT. Additionally, the TC group is anticipated to show greater improvements than the CT group.

#### Theoretical Hypothesis 2 – Stress

It is expected that the interventions including a physical exercise component (DT and TC) will lead to greater reductions in stress levels compared with CT. No significant changes are anticipated in the CT group.

## Materials and Methods

### Study Design

The Muévete conCiencia project is designed as a three-arm, parallel-group randomized controlled trial (DT, TC, CT) with pre- and post-intervention assessments, following a 3 (group) x 2 (time) mixed factorial design with repeated measures. Participants will be randomly allocated in a 1:1:1.

The study will follow the CONSORT (Consolidated Standards of Reporting Trials) guidelines for randomized clinical trials [49] and the guidance for social and psychological interventions [50]. In addition, the protocol has been developed following the SPIRIT (Standard Protocol Items: Recommendations for Interventional Trials) guidelines for study protocols will be applied [51] (see S1 SPIRIT checklist and Fig. 1 SPIRIT schedule of enrolment, interventions, and assessments).

**Fig 1.**
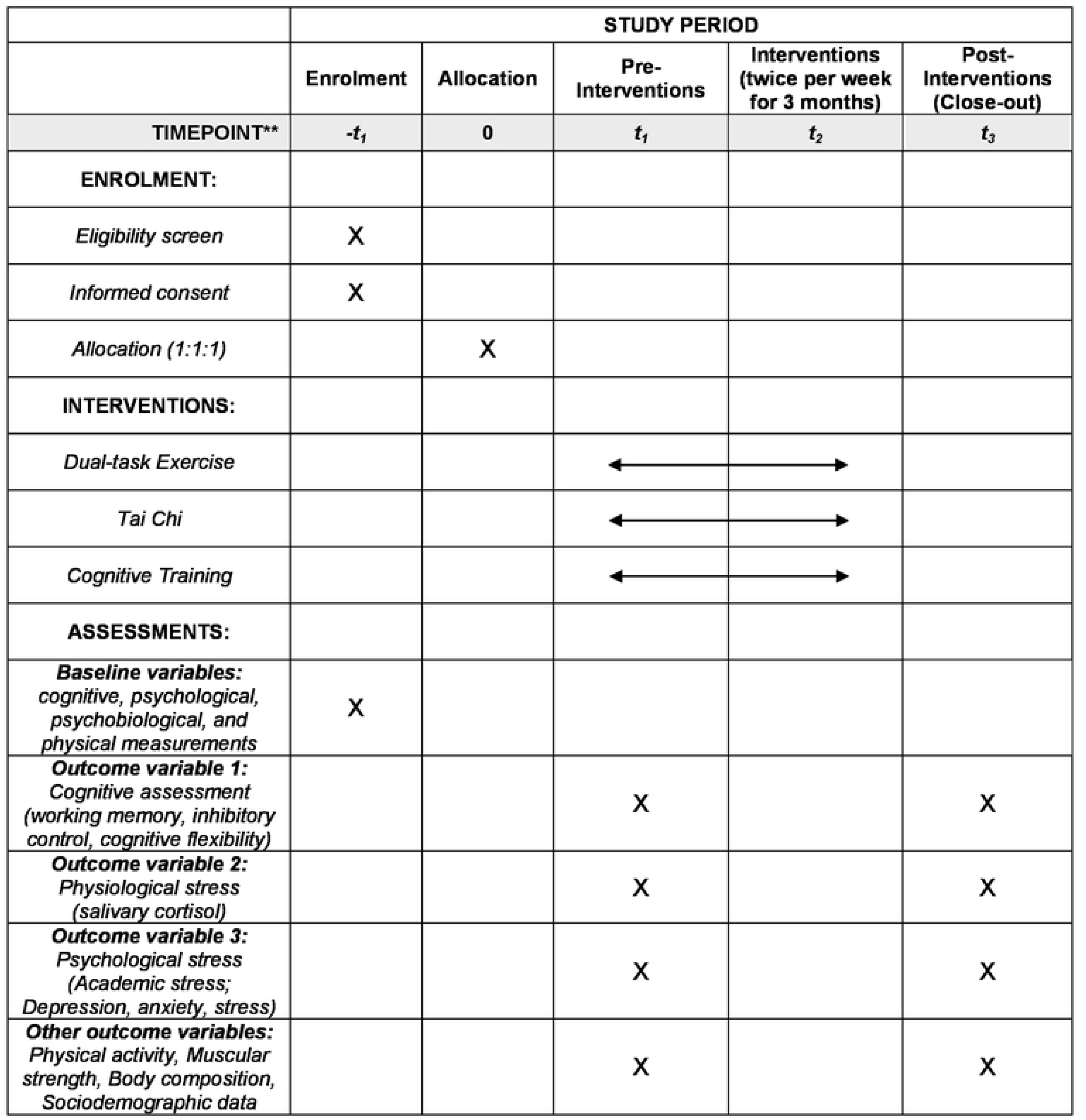
Schedule of enrolment, interventions, and assessments.

The trial has been registered in ClinicalTrials.gov (identifier: NCT07443904) and approved by the Scientific Ethics Committee and the Biosafety Committee of Universidad San Sebastián, Chile. Any protocol amendments will be updated in ClinicalTrials.gov and communicated to regulatory bodies as required.

### Study Setting and Participants

The study will be conducted at Universidad San Sebastián, Concepción campus, Chile. First-year undergraduate students enrolled in health science programs from the 2026 academic cohort will be invited to participate. The interventions will take place in a university gym specifically equipped for physical exercise. All assessments will be conducted in institutional facilities of the university during the regular academic period.

### Sample Size

The required sample size was estimated to detect a statistically significant group × time interaction in a mixed repeated-measures design with three parallel groups (DT, TC, CS) and two assessment points (pre and post). The calculation was performed using G*Power (3.1.9.6.; ANOVA: Repeated measures, within–between interaction). Parameters were set at α = 0.05, power (1−β) = 0.90, and an expected effect size of Cohen’s f = 0.18, corresponding to a small-to-moderate effect according to conventional benchmarks [52]. A correlation among repeated measures of r = 0.50 and a no sphericity correction of ε = 1 were assumed. Under these assumptions, the minimum required sample size was N = 144 (n = 48 per group). To account for an anticipated attrition rate of 10%, the final target sample was increased to N = 159 (n = 53 per group). This sample size is consistent with previous studies evaluating mind–body and physical exercise interventions in university populations [53].

### Recruitment of Participants

Participants will be recruited in March 2026, coinciding with the start of the academic year. Study dissemination will be conducted through multiple institutional channels, including an on-site informational stand, institutional email distribution lists, official university social media platforms, and printed informational posters displayed across campus facilities. These materials will provide general information regarding the study objectives, program duration, session frequency, and assessment procedures.

Students expressing interest will receive detailed information about the study and will be explicitly informed about the research objectives, study procedures, potential benefits and risks, and random allocation to one of the intervention groups. Written informed consent will be obtained from all participants prior to the initiation of any study procedures.

### Inclusion Criteria

Participants will be eligible if they meet all of the following criteria: (1) Aged between 18 and 25 years at the time of enrolment; (2) enrolled as first-year undergraduate students in health science programs at Universidad San Sebastián (Concepción campus) during the 2026 academic cohort; (3) eligible to engage in moderate-to-vigorous physical exercise, as indicated by self-reported absence of contraindications; and (4) available to attend the scheduled intervention sessions and complete pre- and post-intervention assessments.

### Exclusion Criteria

Participants will be excluded if any of the following conditions apply: (1) Presence of diagnosed neurological diseases or severe psychiatric disorders that may interfere with participation in the interventions or compromise the validity of the assessments; (2) medical or musculoskeletal conditions that contraindicate moderate-to-vigorous physical exercise; and (3) regular participation in other structured physical exercise programs during the study period or classification as competitive athletes.

### Randomization

Participants will be allocated to the study groups using a block randomization procedure with stratification by sex (male/female). An allocation ratio of 1:1:1 will be maintained until the target sample of 159 participants is reached (n = 53 per group). The randomization sequence will be generated by an independent researcher not involved in participant recruitment, assessment, or intervention delivery, using a computerized random number generator. Participants will be informed of their assigned group after the baseline evaluation.

Due to the nature of the interventions, participants and instructors cannot be blinded to group allocation. However, outcome assessors conducting the evaluations will remain blinded to group allocation. Unblinding will only occur for post-intervention outcome assessors if a health problem or adverse event requires knowledge of the participant’s assigned intervention. Any instance of unblinding will be documented.

### Baseline assessments

Between March and April 2026, participants will complete baseline (pre-intervention) assessments, including cognitive, psychological, psychobiological, and physical measurements. All assessments will be conducted under standardized conditions by trained evaluators who will be blinded to group allocation. Upon completion of baseline assessments, participants will be randomly assigned to one of the three study groups.

### Interventions

The interventions will be delivered over a 12-week period between April and June 2026, with a frequency of two 60-minute sessions per week. All sessions will be conducted in the university gymnasium under the supervision of qualified instructors responsible for each intervention group (DT, TC, and CT).

Sessions will be scheduled during daytime hours, and alternative time slots will be offered to facilitate participation and promote adherence. Attendance will be systematically recorded throughout the intervention period.

Participants will be allowed to continue their usual daily activities during the trial. However, they will be asked not to initiate structured physical exercise, mind–body training, or cognitive training programs outside the study during the intervention period to avoid potential interference with study outcomes.

If a health problem or adverse event occurs that could compromise the participant’s safety during the sessions, participation may be interrupted. In such cases, the research team will evaluate the situation and may recommend discontinuation of the intervention.

The general structure of the intervention sessions follows the recommendations of the American College of Sports Medicine (ACSM) for exercise prescription [54], which propose an initial preparation phase, a main conditioning phase, and a final recovery phase. This structure was adapted to the specific characteristics of each intervention modality. Exercise intensity and perceived exertion during the intervention will be monitored using the Borg Rating of Perceived Exertion Scale [55], which ranges from 0.5 to 10. A score of 0.5 represents very weak or minimal effort, whereas a score of 10 represents maximal exertion. Adherence to the interventions will be systematically monitored through attendance records, and standardized protocols will be implemented to enhance intervention fidelity [56].

#### Dual-task Exercise (DT)

Each session will include a warm-up phase (10–15 minutes), a main exercise phase (30–35 minutes), and a cool-down phase (10–15 minutes). The warm-up phase will focus on progressive cardiovascular activation, joint mobility, and neuromotor preparation.

The main phase will consist of circuit-based strength, aerobic, and neuromotor exercises integrated with simultaneous cognitive demands, including working memory, inhibitory control, and cognitive flexibility tasks. Cognitive demands will be delivered through verbal instructions and visual stimuli. Participants will perform exercises for 45 seconds followed by 15 seconds of rest, completing two rounds per circuit [57].

Exercise intensity is expected to range from moderate to vigorous (approximately 64–89% of maximum heart rate), monitored using heart rate assessment and the Borg Rating of Perceived Exertion scale, with a gradual and planned progression over the intervention period [58]. During this phase, participants will receive verbal encouragement to promote maximal effort [56].

The cool-down phase will focus on physiological recovery through low-intensity exercises, stretching, and controlled breathing. Sessions will be delivered by an exercise professional with experience in group-based functional training.

#### Tai Chi (TC)

Each TC session will include an opening and centering phase (10–15 minutes), a main Tai Chi practice phase (30–35 minutes), and a final integration and regulation phase (10–15 minutes). The initial phase will emphasize postural alignment, respiratory regulation, and attentional activation through static postures (Wuji), gentle mobility, and conscious breathing.

The main phase will consist of the progressive practice of structured Tai Chi sequences based on a simplified style, integrating slow and continuous movements, weight shifting, limb coordination, postural control, and synchronization of breathing and movement, with an emphasis on mindfulness and motor memory [59].

The final phase will focus on physiological and emotional regulation through gentle closing movements, guided breathing, and conscious relaxation. Exercise intensity will be maintained within low-to-moderate ranges (approximately 55–70% of maximum heart rate), monitored using heart rate and the Borg Rating of Perceived Exertion scale, with gradual and planned progression across weeks [58].

#### Cognitive Training (CT)

Each CT session will include an initial familiarization and task-adjustment phase (5–10 minutes), a main CT phase (40–45 minutes), and a closing and feedback phase (5–10 minutes). The main phase will consist of supervised digital CT using the Graffos software, delivering structured graphomotor tasks.

This intervention involves the simultaneous activation of EF (including sustained and selective attention, working memory, planning, and cognitive flexibility) and fine motor processes. Sessions will be conducted in person under the supervision of a trained professional, who will ensure correct task execution and individual adaptation of task difficulty. The software allows for progressive task programming, provides immediate feedback, and records objective indicators of cognitive performance.

Each session will include structured graphomotor exercises requiring fine motor control, eye–hand coordination, and attentional regulation, thereby promoting functional integration between cognitive and motor processes.

Following completion of the intervention, all participants will undergo post-intervention assessments, replicating the same procedures, instruments, and conditions used at baseline, while maintaining assessor blinding.

### Outcomes

Baseline and post-intervention assessments will be conducted by trained evaluators following standardized procedures. Prior to data collection, all evaluators will receive training to ensure consistency in the administration of cognitive tests, questionnaires, and physical assessments.

Pre- and post-intervention assessments will include cognitive measures (EF), which constitute the primary outcome, the secondary outcomes as psychobiological measures (stress outcomes, including salivary cortisol) and other outcomes are physical measures (anthropometric variables and muscular strength). Data collection forms and assessment protocols will be available from the corresponding author upon request.

### Primary Outcomes Measures

#### Cognitive assessment

Neuropsychological assessments will be administered in a fixed order. The total duration of testing will be approximately 40 minutes, depending on individual performance. Scheduled breaks of 3 minutes will be included to minimize fatigue effects. EF will be assessed using a battery of subtests aligned with the three core dimensions proposed by Miyake et al. [13]. The scores from each prespecified test metric will be converted to Z-scores standardized using the baseline mean and standard deviation.

#### Working memory

Working memory will be assessed using the Working Memory Index (WMI) of the Wechsler Adult Intelligence Scale (WAIS) [60], adapted to the Chilean population in the WAIS-IV version [61]. The WMI includes three subtests: Digit Span (Minimum = 0 - maximum = 48), Arithmetic (Minimum = 0- maximum = 22), and Letter–Number Sequencing (Minimum = 0; maximum = 30), higher scores indicate better working memory.

#### Inhibitory control

Inhibitory control will be assessed using the Stroop Color and Word Test [62], applying normative data from the study by Rivera et al. [63], which integrated results from 11 Latin American countries, including Chile. The test consists of three pages, each containing 100 items arranged randomly in five columns. Participants are given 45 seconds to complete Word (W), Color(C) and Color-Word (CW) as quickly and accurately as possible. Performance is scored as the number of correctly completed items (Minimum = 0; maximum = 100), higher scores indicate better inhibitory control. The interference index will be calculated using the formula CW – [(W × C)/(P + W)], providing an indicator of the individual’s ability to control cognitive interference [63].

#### Cognitive flexibility

Cognitive flexibility will be assessed using the Modified Wisconsin Card Sorting Test (M-WCST) [64]. This test provides indices of the number of categories achieved abstract reasoning, cognitive flexibility, perseverative responses, and total errors. Standardized scores are expressed as T-scores, with a mean of 50 and a standard deviation of 10 (Minimum = 15; maximum = 85), higher scores indicate a better outcome. The instrument has been standardized for individuals aged 6 to 89 years in an Argentine population [66]. Additionally, the Trail Making Test Parts A and B (TMT) using normative data for the Latin American Spanish-speaking adult population, applying the Chile-specific norms reported by Arango-Lasprilla et al. [61]. The TMT assesses the ability to alternate between mental sets and manage divided attention demands. The score is the completion time for each part of the TMT-A (Minimum = 0; maximum = 100) and TMT-B (Minimum = 0; maximum = 300). Lower completion time indicates better performance.

### Secondary Outcomes Measures

#### Physiological stress: Salivary cortisol

Salivary cortisol levels reflect physiological stress associated with HPA axis activity. Changes in physiological stress in response to the interventions will be assessed non-invasively through salivary cortisol determination [67,68], a validated approach for the study of academic stress in educational contexts [69]. Cortisol concentrations will be expressed in µg/dL. The analytical measurement range of the assay is approximately 0.012–3.0 µg/dL. Higher values indicate greater physiological stress.

#### Saliva sample collection

Saliva samples will be collected at the Medical Technology laboratories of Universidad San Sebastián using the passive drool method, which is currently considered the gold standard for salivary biomarker assessment due to its ability to preserve sample integrity and avoid analytical interference [70].

To allow duplicate cortisol determination (approximately 1–2 mL), participants will be scheduled for sample collection at 08:00 a.m. Participants will accumulate saliva in the floor of the mouth and transfer it into sterile 1.5 mL polypropylene tubes. To minimize pre-analytical errors, participants will be instructed to refrain from food intake, beverages (except water), smoking, and oral hygiene procedures for at least 30 minutes prior to sample collection. Additionally, participants will be asked to record their wake-up time on the day of sampling.

Following collection, samples will be processed by laboratory personnel and centrifuged to remove cellular debris and mucins. The supernatant will be stored at −20 °C until final analysis.

#### Salivary cortisol determination

Cortisol concentration will be quantified using a competitive enzyme-linked immunosorbent assay (ELISA) with the commercial kit EIAHCOR (Invitrogen). Prior to analysis, saliva samples will be diluted at a minimum ratio of 1:4 using the provided 1× Assay Buffer. The assay will be performed according to the manufacturer’s instructions. Optical density will be measured at 450 nm, and cortisol concentrations will be calculated using an eight-point standard curve.

### Psychological stress: Self-report measures

#### Academic stress

Academic stress will be assessed using the Academic Stress Inventory [71], a 33-item scale evaluating three dimensions: stressors, coping strategies, and stress responses, which include physical, psychological, and behavioral reactions. Responses are recorded on a 5-point Likert scale ranging from 1 (“never”) to 5 (“always”) the score (Minimum = 33; maximum = 165). Lower completion time indicates better performance. In the Chilean context, the scale has demonstrated adequate reliability, with Cronbach’s alpha and McDonald’s omega values of 0.78 for the Stressor factor, and alpha values of 0.89 for physical and psychological reactions and 0.85 for socio-behavioral reactions [72]. The scale will be administered online via a QR code and monitored to ensure completion during the two-week assessment period.

#### Depression, anxiety, and stress symptoms

Symptoms of depression, anxiety, and stress will be assessed using the Depression Anxiety Stress Scales (DASS-21), adapted for the Chilean population by Antúnez & Vinet [73]. The scale consists of 21 items rated on a Likert scale from 0 (“did not apply to me at all”) to 5 (“applied to me very much or most of the time”), reflecting the extent to which each statement describes the participant’s experiences over the previous week (Minimum = 0; maximum = 126). Higher scores indicate more global symptoms. The DASS-21 is a brief self-report measure with well-established psychometric properties in university student populations. The scale will be administered online via a QR code and monitored to ensure completion during the two-week assessment period.

### Other Outcomes Measures

#### Physical activity levels

Physical activity levels will be assessed using the short form of the International Physical Activity Questionnaire (IPAQ-SF), validated for the Chilean population [74]. The instrument provides information on physical activity performed during the previous seven days, including light, moderate, and vigorous intensity activity, frequency (days/week), and duration (minutes/day). The questionnaire consists of seven items; for example, participants are asked how many days they engaged in vigorous activities such as lifting heavy objects, intensive sports practice, running, or fast cycling. Responses range from 0 to 7 days, and reported minutes are summed accordingly. The score will be expressed as MET-minutes per week where higher scores indicate higher levels of physical activity.

#### Muscular strength

Muscular strength will be assessed using a handgrip dynamometer following standardized protocols. Three trials will be performed for each hand, and the highest value (kg/force) will be recorded. Higher values indicate greater muscle strength. Handgrip strength is a marker of physical function and has been associated with functional performance and health outcomes in adults [75].

#### Body composition

Body composition will be assessed using bioelectrical impedance analysis to determine body fat percentage and fat-free mass percentage. A standardized protocol will be applied to ensure measurement reliability, including a minimum of 6 hours of fasting, avoidance of alcohol and physical exercise for 12 hours prior to assessment, bladder voiding before measurement, and exclusion of measurements during menstruation [75,76]. Outcomes include Body weight (kg), Height (meters), body fat percentage (%), and fat-free mass percentage (%) will be recorded. Body mass index (BMI) will be calculated as weight divided by height squared [BMI = weight (kg)/height² (m²)] [76].

#### Sociodemographic variables

Sociodemographic variables will include sex and age of the participants.

##### Intervention adherence

Adherence to the intervention program will be assessed by recording attendance at scheduled sessions. An adherence percentage will be calculated as (number of attended sessions/24 scheduled sessions) × 100.

##### Conceptual synthesis and scope of the program

The mind–body program adopts an integrative approach that targets physiological, cognitive, and emotional regulation through conscious movement. All sessions are delivered in a non-competitive and supportive group environment, encouraging body awareness, self-observation, and emotional regulation. Exercise intensity is individually monitored using heart rate and perceived exertion, ensuring a safe and progressive intervention aligned with the study objectives.

##### Statistical Analysis

Missing data will be handled using multiple imputation methods [78]. Descriptive analyses will be conducted using measures of central tendency and dispersion for continuous variables, and absolute and relative frequencies for categorical variables. Data normality will be assessed using the Shapiro–Wilk or Kolmogorov–Smirnov tests, as appropriate. A two-step transformation approach will be applied to non-normally distributed variables [79]. Homoscedasticity will be examined using Levene’s test. A two-way repeated-measures analysis of variance (ANOVA) will be performed to evaluate intervention effects over time. Fixed effects will include intervention group DT, TC and CT, time (pre- and post-intervention), and their interaction (time × group). When statistically significant effects are identified, post hoc comparisons with Bonferroni adjustment will be conducted. Effect sizes will be calculated using Cohen’s d and interpreted as follows: <0.20 trivial, ≥0.20 and ≤0.49 small, ≥0.50 and ≤0.79 moderate, and ≥0.80 large [80]. Internal consistency of the scales will be evaluated using Cronbach’s alpha and McDonald’s omega coefficients. All statistical analyses will be performed using JASP statistical software (version 0.95.4), with the level of statistical significance set at p < 0.05.

#### Data monitoring

A formal Data Monitoring Committee (DMC) will not be established due to the low-risk nature of the interventions, which involve supervised physical exercise, Tai Chi, and cognitive training activities. Study oversight will be conducted by the principal investigator and the research team, who are independent from the study sponsor and funder.

No interim analyses or formal stopping guidelines are planned due to the minimal risk and short duration of the trial. Trial conduct will be periodically reviewed by the research team to ensure protocol adherence, data quality, and participant safety.

#### Ethical Considerations

The study will be conducted in accordance with the ethical principles for research involving human participants established in the Declaration of Helsinki [77]. The protocol was reviewed and approved by the Scientific Ethics Committee and the Biosafety Committee of Universidad San Sebastián (approval date: May 16, 2025; approval code No. 57-25). The program will be implemented in compliance with institutional biosafety standards and requirements (CB-25-018).

Ethical safeguards will be maintained in the handling of participant information, which will be treated as confidential. Data collection procedures will not pose risks to participants’ integrity. Participation will be voluntary, and participants will be free to withdraw from the study at any time without consequences. Data generated during the study will be made available upon request following publication of the primary study results. The shared dataset will include the data dictionary necessary to interpret the variables used in the study.

## Discussion

The implementation of randomized clinical trials in first-year university students entails relevant methodological and operational challenges, including logistical issues related to adherence to scheduled activities, coordination of interventions with academic workload, and potential interference from academic assessment periods. In addition, the collection of salivary biomarkers requires strict adherence to sampling schedules and instructions, which may affect data quality if not carefully controlled. To mitigate these risks, systematic reminders, WhatsApp communication groups, student research assistants, flexible scheduling, and a structured induction process for the assessment and intervention teams will be implemented.

All three interventions are expected to produce improvements in EF, with greater gains anticipated in the group receiving higher-intensity physical exercise. Regarding stress outcomes, including psychological stress and salivary cortisol, reductions are expected in both physical exercise groups DT and TC compared with the CT group. Additionally, potential sex-related differences in intervention effects will be explored.

A primary limitation of this study is that it will be conducted at a single Chilean university, which may limit the generalizability of the findings to other institutional contexts. Study results will be disseminated through publications in peer-reviewed scientific journals and presentations at national and international conferences, with the aim of informing and promoting health promotion programs within university settings.

Any protocol amendments will require approval from the Ethics Committee of Universidad San Sebastián and will be reported in future publications in accordance with CONSORT guidelines for randomized clinical trials. The study may be terminated prematurely in the event of unforeseen risks to participant safety, insurmountable recruitment difficulties, or administrative or institutional constraints. In such cases, the ethics committee, the sponsoring institution, and, when appropriate, the participants will be promptly informed, ensuring confidentiality and participant welfare.

In conclusion, this protocol describes a pilot randomized controlled trial designed to evaluate the effects of different modalities of physical exercise, mind–body practice, and cognitive training on university students. Its implementation aims to contribute evidence from a Latin American context and to inform future interventions promoting health and well-being in higher education.

## Data Availability

All relevant data from this study will be made available upon study completion.

## Author Contributions

MRV: Conceptualization; Methodology; Investigation; Project Administration; Supervision; Funding Acquisition; Writing – Original Draft Preparation; Writing – Review & Editing.

CP: Investigation; Supervision of physical assessments and exercise-related measurements; Writing – Review & Editing.

CB: Investigation; Supervision of physical assessments and exercise-related measurements; Writing – Review & Editing.

CL: Investigation; Supervision of salivary cortisol collection and laboratory procedures; Writing – Review & Editing.

DRM: Methodology; Investigation; Supervision; Writing – Review & Editing.

AK: Methodology; Program Design; Writing – Original Draft Preparation; Writing – Review & Editing.

DP-O: Methodology; Writing – Review & Editing.

SR: Methodology; Writing – Writing – Review & Editing. IdO:Methodology; Writing – Writing – Review & Editing.

CP-D: Methodology; Program Design; Writing – Original Draft Preparation; Writing – Review & Editing.

## Acknowledgments

The authors gratefully acknowledge B-Graffos International S.L. for the free provision of the Graffos software, which was instrumental in supporting this research.

---

SPIRIT 2025 checklist of items to address in a randomized trial protocol
| Section / Topic | No | SPIRIT 2025 checklist item description | Reported on page no. |
| --- | --- | --- | --- |
| **Administrative information** |  |  |  |
| Title and structured summary | 1a | Title stating the trial design, population, and interventions, with identification as a protocol | 1 |
|  | 1b | Structured summary of trial design and methods, including items from the World Health Organization Trial Registration Data Set | 2 |
| Protocol version | 2 | Version date and identifier | 2 |
| Roles and responsibilities | 3a | Names, affiliations, and roles of protocol contributors | 1, 16 |
|  | 3b | Name and contact information for the trial sponsor | 1 |
|  | 3c | Role of trial sponsor and funders in design, conduct, analysis, and reporting of trial; including any authority over these activities | 16 |
|  | 3d | Composition, roles, and responsibilities of the coordinating site, steering committee, endpoint adjudication committee, data management team, and other individuals or groups overseeing the trial, if applicable | 7 |
| **Open science** |  |  |  |
| Trial registration | 4 | Name of trial registry, identifying number (with URL), and date of registration. If not yet registered, name of intended registry | 7 |
| Protocol and statistical analysis plan | 5 | Where the trial protocol and statistical analysis plan can be accessed | 6, 14 |
| Data sharing | 6 | Where and how the individual de-identified participant data (including data dictionary), statistical code, and any other materials will be accessible | 14 |
| Funding and conflicts of interest | 7a | Sources of funding and other support (e.g., supply of drugs) | 16 |
|  | 7b | Financial and other conflicts of interest for principal investigators and steering committee members | 16 |
| Dissemination policy | 8 | Plans to communicate trial results to participants, healthcare professionals, the public, and other relevant groups (e.g., reporting in trial registry, plain language summary, publication) | 15 |
| **Introduction** |  |  |  |
| Background and rationale | 9a | Scientific background and rationale, including summary of relevant studies (published and unpublished) examining benefits and harms for each intervention | 3-5 |
|  | 9b | Explanation for choice of comparator | 4-5 |
| Objectives | 10 | Specific objectives related to benefits and harms | 5 |
| **Methods: Patient and public involvement, trial design** |  |  |  |
| Patient and public involvement | 11 | Details of, or plans for, patient or public involvement in the design, conduct, and reporting of the trial | 6 |
| Trial design | 12 | Description of trial design including type of trial (e.g., parallel group, crossover), allocation ratio, and framework (e.g., superiority, equivalence, non-inferiority, exploratory) | 6 |

| Methods: Participants, interventions, and outcomes |  |  |  |
| --- | --- | --- | --- |
| Trial setting | 13 | Settings (e.g., community, hospital) and locations (e.g., countries, sites) where the trial will be conducted | 6 |
| Eligibility criteria | 14a | Eligibility criteria for participants | 7 |
|  | 14b | If applicable, eligibility criteria for sites and for individuals who will deliver the interventions (e.g., surgeons, physiotherapists) | 7 |
| Intervention and comparator | 15a | Intervention and comparator with sufficient details to allow replication including how, when, and by whom they will be administered. If relevant, where additional materials describing the intervention and comparator (e.g., intervention manual) can be accessed | 8 |
|  | 15b | Criteria for discontinuing or modifying allocated intervention/comparator for a trial participant (e.g., drug dose change in response to harms, participant request, or improving/worsening disease) | 8 |
|  | 15c | Strategies to improve adherence to intervention/comparator protocols, if applicable, and any procedures for monitoring adherence (e.g., drug tablet return, sessions attended) | 8 |
|  | 15d | Concomitant care that is permitted or prohibited during the trial | 8 |
| Outcomes | 16 | Primary and secondary outcomes, including the specific measurement variable (e.g., systolic blood pressure), analysis metric (e.g., change from baseline, final value, time to event), method of aggregation (e.g., median, proportion), and time point for each outcome | 11-13 |
| Harms | 17 | How harms are defined and will be assessed (e.g., systematically, non-systematically) | 6 |
| Participant timeline | 18 | Time schedule of enrollment, interventions (including any run-ins and washouts), assessments, and visits for participants. A schematic diagram is highly recommended (see Figure) | 6 |
| Sample size | 19 | How sample size was determined, including all assumptions supporting the sample size calculation | 6 |
| Recruitment | 20 | Strategies for achieving adequate participant enrollment to reach target sample size | 7 |
| Methods: Assignment of interventions |  |  |  |
| Randomization: |  |  |  |
| Sequence generation | 21a | Who will generate the random allocation sequence and the method used | 8 |
|  | 21b | Type of randomization (simple or restricted) and details of any factors for stratification. To reduce predictability of a random sequence, other details of any planned restriction (e.g., blocking) should be provided in a separate document that is unavailable to those who enroll participants or assign interventions | 8 |
| Allocation concealment mechanism | 22 | Mechanism used to implement the random allocation sequence (e.g., central computer/telephone; sequentially numbered, opaque, sealed containers), describing any steps to conceal the sequence until interventions are assigned | 8 |
| Implementation | 23 | Whether the personnel who will enroll and those who will assign participants to the interventions will have access to the random allocation sequence | 8 |
| Blinding | 24a | Who will be blinded after assignment to interventions (e.g., participants, care providers, Unblinding will only occur for post-intervention outcome assessors if a health problem or adverse event requires knowledge of the participant's assigned intervention. Any instance of unblinding will be documented. , data analysts) | 8 |
|  | 24b | If blinded, how blinding will be achieved and description of the similarity of interventions | 8 |
|  | 24c | If blinded, circumstances under which unblinding is permissible, and procedure for revealing a participant's allocated intervention during the trial | 8 |
| **Methods: Data collection, management, and analysis** |  |  |  |
| Data collection methods | 25a | Plans for assessment and collection of trial data, including any related processes to promote data quality (e.g., duplicate measurements, training of assessors) and a description of trial instruments (e.g., questionnaires, laboratory tests) along with their reliability and validity, if known. Reference to where data collection forms can be accessed, if not in the protocol | 10-15 |
|  | 25b | Plans to promote participant retention and complete follow-up, including list of any outcome data to be collected for participants who discontinue or deviate from intervention protocols | 14 |
| Data management | 26 | Plans for data entry, coding, security, and storage, including any related processes to promote data quality (e.g., double data entry; range checks for data values). Reference to where details of data management procedures can be accessed, if not in the protocol |  |
| Statistical methods | 27a | Statistical methods used to compare groups for primary and secondary outcomes, including harms | 14 |
|  | 27b | Definition of who will be included in each analysis (e.g., all randomized participants), and in which group | 14 |
|  | 27c | How missing data will be handled in the analysis | 14 |
|  | 27d | Methods for any additional analyses (e.g., subgroup and sensitivity analyses) | 14 |
| **Methods: Monitoring** |  |  |  |
| Data monitoring committee | 28a | Composition of data monitoring committee (DMC); summary of its role and reporting structure; statement of whether it is independent from the sponsor and funder; conflicts of interest and reference to where further details about its charter can be found, if not in the protocol. Alternatively, an explanation of why a DMC is not needed | 15 |
|  | 28b | Explanation of any interim analyses and stopping guidelines, including who will have access to these interim results and make the final decision to terminate the trial | 15 |
| Trial monitoring | 29 | Frequency and procedures for monitoring trial conduct. If there is no monitoring, give explanation | 15 |
| **Ethics** |  |  |  |
| Research ethics approval | 30 | Plans for seeking research ethics committee/institutional review board approval | 15 |
| Protocol amendments | 31 | Plans for communicating important protocol modifications to relevant parties | 6 |
| Consent or assent | 32a | Who will obtain informed consent or assent from potential trial participants or authorized proxies, and how | 7 |
|  | 32b | Additional consent provisions for collection and use of participant data and biological specimens in ancillary studies, if applicable | 7 |
| Confidentiality | 33 | How personal information about potential and enrolled participants will be collected, shared, and maintained in order to protect confidentiality before, during, and after the trial | 15 |
| Ancillary and post-trial care | 34 | Provisions, if any, for ancillary and post-trial care, and for compensation to those who suffer harm from trial participation | 15 |

© 2025 Chan A-W et al. This is an Open Access article distributed under the terms of the Creative Commons Attribution License (https://creativecommons.org/licenses/by/4.0/), which permits unrestricted use, distribution, and reproduction in any medium, provided the original work is properly cited.

